# Impact of non-pharmaceutical interventions on documented cases of COVID-19

**DOI:** 10.1101/2020.04.16.20062141

**Authors:** Nicolas Banholzer, Eva van Weenen, Bernhard Kratzwald, Arne Seeliger, Daniel Tschernutter, Pierluigi Bottrighi, Alberto Cenedese, Joan Puig Salles, Werner Vach, Stefan Feuerriegel

## Abstract

**Background:** The novel coronavirus (SARS-CoV-2) has rapidly evolved into a global epidemic. To control its spread, countries have implemented non-pharmaceutical interventions (NPIs), such as school or border closures, while others have even enforced complete lockdowns. Here we study the impact of NPIs in reducing documented cases of COVID-19. Documented case numbers are selected because they are essential for decision-makers in the area of health-policy when monitoring and evaluating current control mechanisms.

**Methods:** We empirically estimate the relative reduction in the number of new cases attributed to each NPI. A cross-country analysis is performed using documented cases through April 15, 2020 from *n* = 20 countries (i.e., the United States, Canada, Australia, the EU-15 countries, Norway, and Switzerland).

**Results:** As of April 15, venue closures were associated with a reduction in the number of new cases by 36 % (95% credible interval [CrI] 20–48 %), closely followed by gathering bans (34 %; 95% CrI 21–45 %), border closures (31 %; 95% CrI 19–42 %), and work bans on non-essential business activities (31 %; 95% CrI 16–44 %). Event bans lead to a slightly less pronounced reduction (23 %; 95% CrI 8–35 %). School closures (8 %; 95% CrI 0–23 %) and lockdowns (5 %; 95% CrI 0–14 %) appeared to be the least effective among the NPIs considered in this analysis.

**Conclusions:** With this cross-country analysis, we provide early estimates regarding the impact of different NPIs for controlling the COVID-19 epidemic. These findings are relevant for evaluating current health-policies.

## 1. Introduction

The novel coronavirus (SARS-CoV-2) has evolved into a global epidemic [1]. The coronavirus was first identified in Wuhan, China [2, 3, 4, 5], but quickly spread across China and the rest of the world [6]. As of April 21, 2020, the total number of confirmed cases of COVID-19, the disease caused by the coronavirus, exceeded more than 2.3 million worldwide. Efforts to control the spread of SARS-CoV-2 focused on non-pharmaceutical interventions (NPIs). These represent public health-policy measures that were intended to diminish transmission rates and, to this end, aimed at reducing person-to-person contacts via so-called social distancing [7]. Examples include school closures, travel restrictions, or even complete lockdowns.

NPIs have been linked to the spread of SARS-CoV-2 primarily within or from a single country [8, 9, 10, 11, 12, 13, 14, 15, 16, 17, 18, 19], mostly focusing on the novel coronavirus outbreak in China. The majority of findings are based on transmission models where epidemiological parameters are informed by previous studies or corroborated via simulation. One study analyzed the impact of NPIs using data from different countries and found that the NPIs considered were together effective in reducing transmission rates [20], using the link between true unobserved infections and deaths. Here we focus on the relative impact of individual NPIs in reducing the number of documented cases. This is yet unknown, although documented COVID-19 cases are essential for decision-makers in health-policy when monitoring and evaluating the impact of NPIs (e.g., controlling the outbreak in a way that surge capacities are not exceeded).

We empirically estimated how NPIs were associated with changes in documented COVID-19 cases through April 15, 2020 across *n* = 20 countries (i.e., the United States, Canada, Australia, the EU-15 countries, Norway, and Switzerland). This amounts to ∼1.6 million documented COVID-19 cases. Our estimation model allowed us to quantify the relative reduction in the number of new cases attributable to each NPI, accounting for the time delay until NPIs became effective, potential effects from the day of the week, and for differences in the speed of disease spread from country to country.

## 2. Methods

### 2.1. Data on documented COVID-19 cases

A cross-country analysis for *n* = 20 countries was performed. The sample comprises the United States, Canada, Australia, the EU-15 countries (Austria, Belgium, Denmark, Finland, France, Germany, Greece, Ireland, Italy, Luxembourg, the Netherlands, Portugal, Spain, Sweden, and the United Kingdom), Norway, and Switzerland. Our selection of countries defines a sample that followed a similar and comparable overall strategy in controlling the COVID-19 outbreak and shares a common cultural background. In particular, Asian countries were excluded despite the availability of excellent data, as these countries have often responded quite differently based on their experience with previous pandemics such as the 2013 SARS-CoV-1 outbreak. Nevertheless, the sample entailed considerable variation across countries, as some countries have been affected severely, while others responded early as part of their mitigation strategy. Overall, the sample covers a population of ∼0.8 billion people. SARS-CoV-2 infection figures for each country were obtained from the Johns Hopkins Coronavirus Resource Center, which was developed for real-time tracking of reported cases of the coronavirus disease 2019 (COVID-19) and directly aggregates cases recorded by local authorities in order to overcome time delays from alternative reporting bodies [21]. Hence, these numbers are supposed to account for all COVID-19 cases identified on a specific day. Case numbers were collected through April 15, 2020. In total, our sample comprises ∼1.6 million cases.

### 2.2. Data on non-pharmaceutical interventions

NPIs were systematically obtained from government resources and news outlets before being classified into seven categories (see Tbl. 1): (1) school closures, (2) border closures, (3) public event bans, (4) gathering bans, (5) venue closures (e.g., shops, bars, restaurants, and venues for other recreational activities), (6) lockdowns prohibiting public movements without valid reason, and (7) work bans on non-essential business activities. Note that “border closures” represents a measure that was fairly severe in restricting international travel. Lockdowns always also implied the NPIs event bans, gathering bans and venue closures, if these have not been explicitly implemented earlier. The timing was encoded such that it refers to the first day an NPI went into effect. We only considered NPIs that were implemented throughout a country or in at least two thirds of its regions. Sources and details on data collection for non-pharmaceutical interventions are provided in Appendix H.

**Table 1:** List of non-pharmaceutical interventions (NPIs), their definitions, and the number of countries that implemented the respective NPI.

| NPI | Definition | Freq. |
| --- | --- | --- |
| School closure | Closure of schools (for primary schools) | 18 |
| Border closure | Closure of national borders for individuals | 11 |
| Event ban | Cancellation of mass gatherings (i.e., 50 people or more) | 20 |
| Gathering ban | Prohibition of small gatherings in public or private spaces of people not from the same household | 19 |
| Venue closure | Closure of venues for recreational activities and/or closure of shops, bars, and restaurants | 19 |
| Lockdown | Prohibition of movement without valid reason (e.g., restricting mobility except to/from work, local supermarkets, and pharmacies) | 11 |
| Work ban | Closure of non-essential business activities (i.e., all businesses except supermarkets, food suppliers, and pharmacies), thus prohibiting corresponding mobility | 6 |

Overall, the number and timing of implemented NPIs varied across countries (Fig. 1). Within the period of our analysis, school closures were implemented by 18 out of the *n* = 20, lockdowns by 11, and work bans by 6. Rising numbers of documented cases prompted more severe NPIs.

**Fig. 1.**
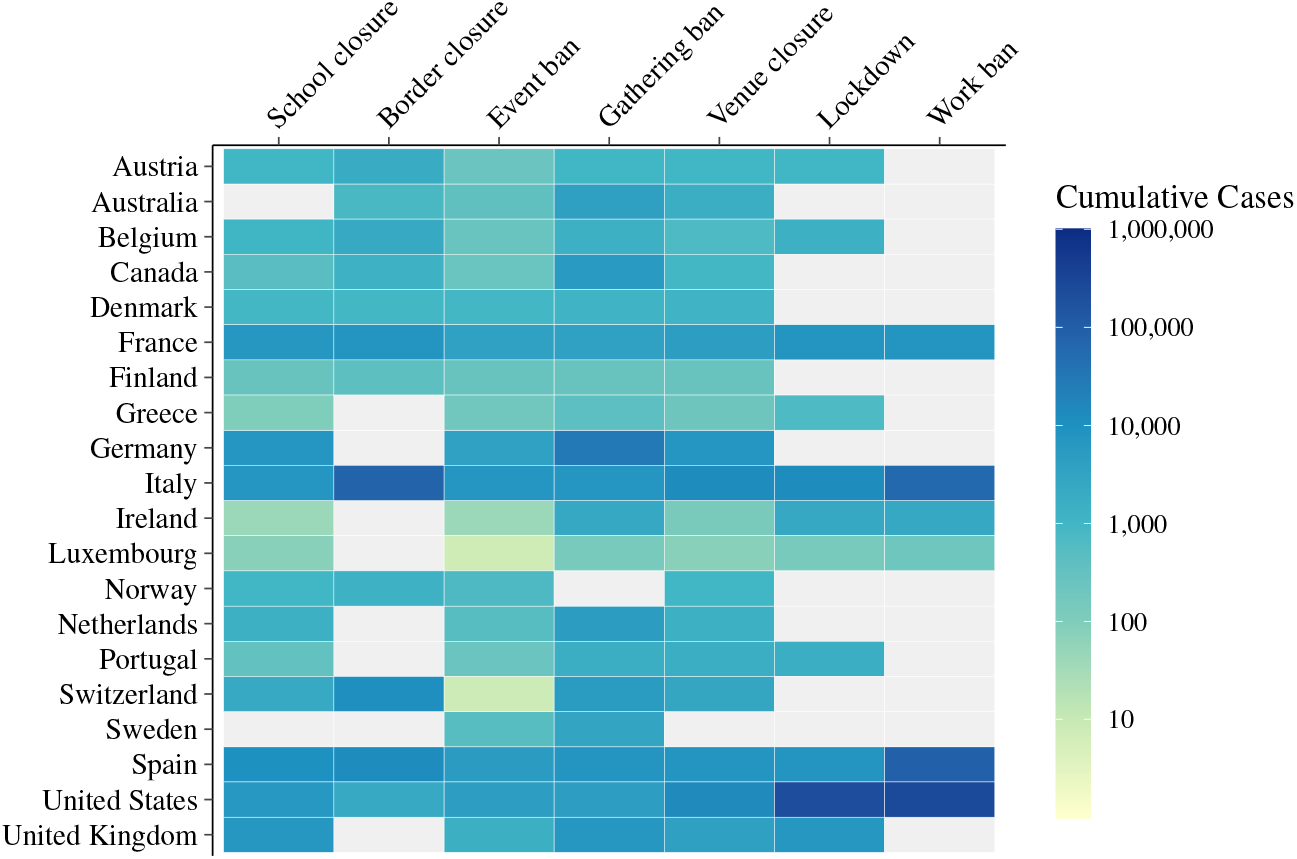
Heatmap showing the cumulative number of documented COVID-19 cases for each country until a non-pharmaceutical intervention (NPI) was implemented. By reporting the cumulative number of cases, the heatmap highlights the ordering of NPIs within countries. If an NPI was not implemented by a country, it is colored gray. A heatmap showing the cumulative number of cases relative to the country’s population is shown in Appendix Fig. 1.

Our outcome of interest was the number of new documented COVID-19 cases per day. This choice reflects that the true number of new cases was unknown and, therefore, the documented numbers served as the basis on which NPIs were monitored and evaluated by decision-makers in health-policy. In particular, case numbers had to be controlled to an extent, so that surge capacities in critical care were not exceeded at the peak of the epidemic [22]. Fig. 2 shows the number of new cases over time with the implemented NPIs indicated. Further descriptive statistics of our data are shown in Appendix A.

**Fig. 2.**
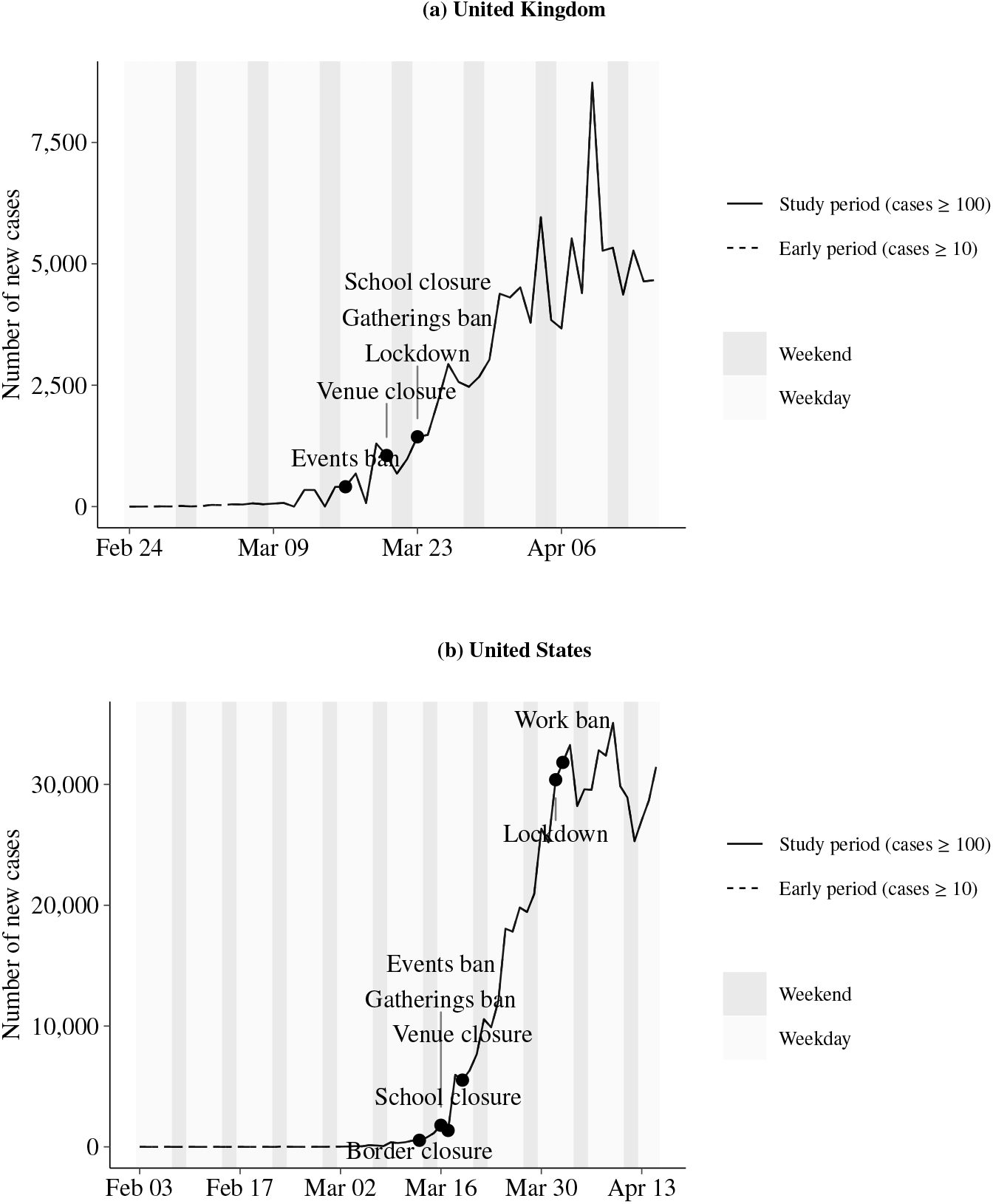
Temporal development of the observed (documented) number of new cases by country (*n* = 20). The points at which non-pharmaceutical interventions (NPIs) were implemented are annotated. The solid line refers to the study period, starting from a cumulative case count of 100. To show the timing of NPIs potentially implemented before that period, the dashed line refers to the early period, starting from a cumulative case count of 10.

### 2.3. Statistical analysis

Daily number of new cases of COVID-19 were linked to the NPIs that were already implemented. It was assumed that NPIs could only influence the number of new cases with a delay of *t*_0_ = 7 days. This choice of the delay considered the following factors. First, prior research on SARS-CoV-2 transmissions indicated an incubation period of around 5 days [3]. Second, as testing was often performed only in response to symptoms and positive results may have been reported with some delay, a further overall delay of about two days was thus not unlikely. Third, behavioral responses could have varied with people adjusting their behavior with a certain delay or even before the NPI was issued. Given the overall uncertainty in the delay at which NPIs took effect, the sensitivity of our results to the chosen time delay of *t*_0_ = 7 days was studied as part of a sensitivity analysis.

The daily number of new cases were related to the actual number of cumulative cases, such that the estimated effect for each NPI quantified the relative reduction in new cases. The model considered day-of-the-week effects as the frequency of testing may have depended on the day of the week and reporting may have been delayed to a higher degree during weekends. The background rate of new cases was allowed to vary across countries as this rate depends on the (unknown) age composition of the true cases, the population density, and other country-specific factors.

The very early phase up to 100 documented cases was excluded from the analysis as countries still had to establish their documentation practices in the early phase. However, the number of cases at the start was varied as part of a sensitivity analysis. The effects of all NPIs were estimated simultaneously in a Bayesian hierarchical model assuming a negative binomial distribution for the daily number of new cases. Details about model specification and estimation procedure are given in Appendices B and C. In the results, we report the posterior mean and 80% and 95% credibility intervals for the relative reductions in the number of new daily cases for each NPI.

## 3. Results

### 3.1. Impact of non-pharmaceutical interventions

Using data from the early stages of the outbreak until April 15, 2020, the relative reduction in the number of new cases was estimated for each NPI (Fig. 3, detailed estimation results are to be found in Appendix D). Venue closures were associated with the highest reduction in the number of new cases (36 %; 95% CrI 20–48 %). The mean reduction was slightly lower for gathering bans (34 %; 95% CrI 21–45 %), border closures (31 %; 95% CrI 19–42 %), and work bans on non-essential business activities (31 %; 95% CrI 16–44 %). Event bans were associated with a slightly less pronounced reduction (23 %; 95% CrI 8–35 %), whereas school closures (8 %; 95% CrI 0–23 %) and lockdowns (5 %; 95% CrI 0–14 %) appeared to be the least effective among the NPIs considered in this analysis.

**Fig. 3.**
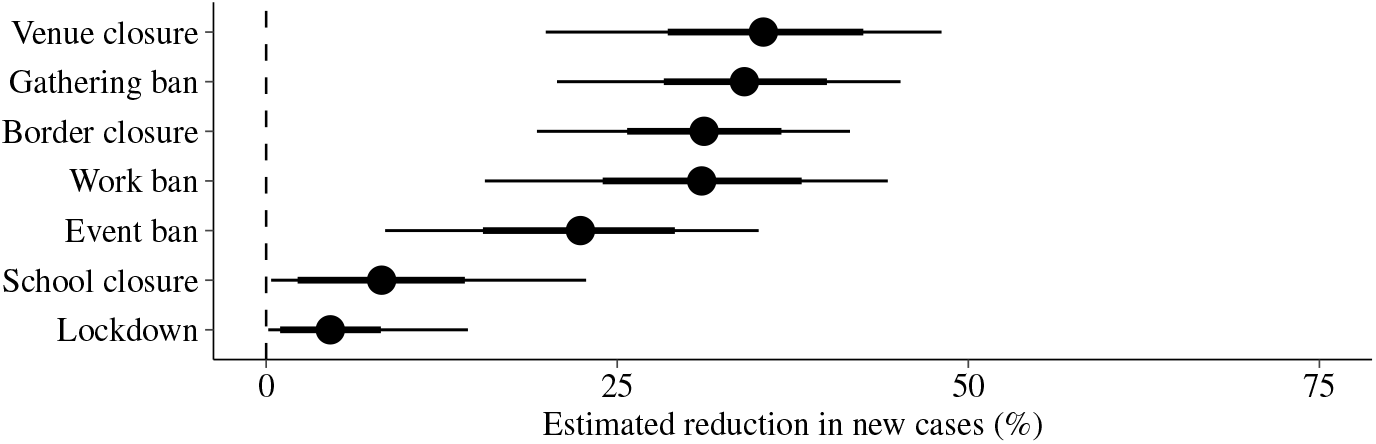
Estimated reduction (posterior mean as dots with 80% and 95% credible interval as thick and thin lines, respectively) in the number of new cases (in %) for each non-pharmaceutical intervention (NPI).

For a precise ranking of the impact of NPIs, the range of credible effects has to be considered. Thereby, venue closures, gathering bans, border closures, and work bans showed similar effect sizes with a magnitude of the relative reduction above 15 %. With respect to event bans, an effect close to 0 % was precluded but the range of credible effects included a moderate relative reduction by 10 % up to much larger reductions. The posterior distributions of school closures and lockdowns focused on small effects with a magnitude of 0 % to 10 %, but for school closures a 20 % reduction was still credible.

Fig. 4 directly depicts the posterior distribution of the ranking of effects. For each of the four NPIs with the highest effect estimates we could be at least 85 % sure that they were among the four most effective NPIs. Conversely, for school closures and lockdowns, we could be at least 85 % sure that they were among the two least effective NPIs.

**Fig. 4.**
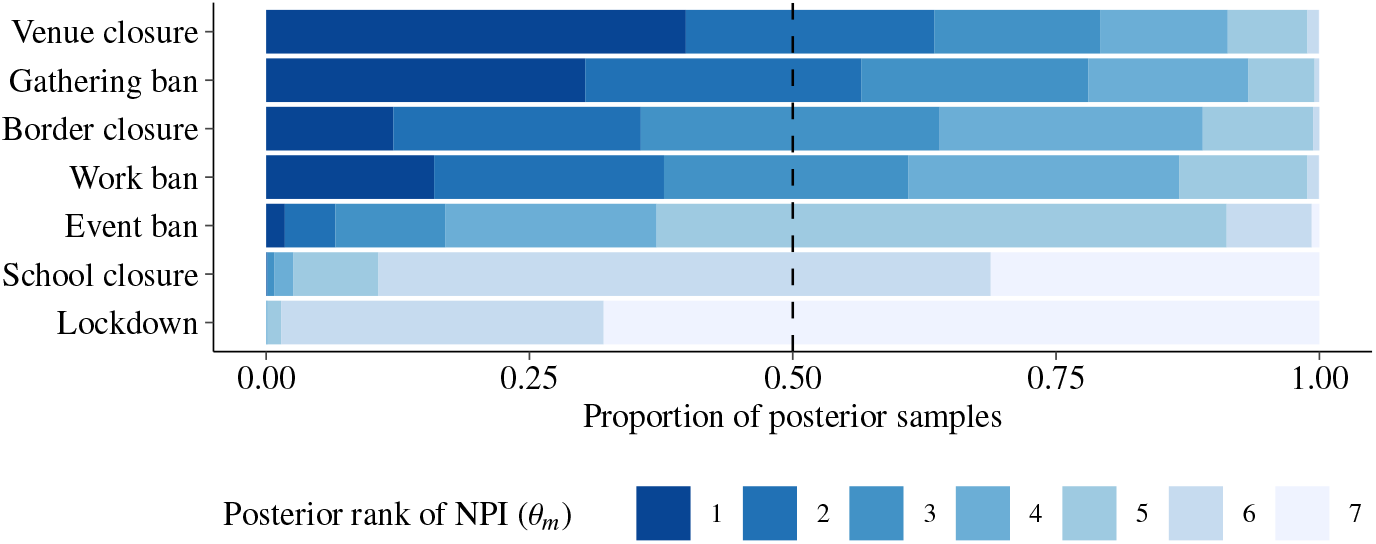
Posterior distribution of ranking non-pharmaceutical interventions (NPIs) by their effect. Shown is the proportion of posterior samples (4,000 in total) for each rank of the NPI parameter (*θ*_*m*_). Highest reductions obtained rank 1 and lowest reductions rank 7.

### 3.2. Sensitivity analysis

Results from the sensitivity analysis are reported in Appendix E. A small variation in the estimated effect of NPIs was observed, in particular for work bans, representing the NPI with the lowest rate of implementation. A model comparison (Appendix F) further suggested that an influence function with a longer time delay and/or a first-order spline would be beneficial, but the estimated reduction of each NPI remained qualitatively similar in these alternative model specifications.

### 3.3. Illustrated hypothetical outcomes

The impact of different NPIs on new COVID-19 cases in single countries is illustrated in Fig. 5. For this purpose, the effect estimate of the corresponding NPI was used to predict the hypothetical number of new cases as if the NPI was not implemented. In this hypothetical setting, the number of new cases is subject to considerable growth. In contrast, the observed (documented) count remains well below the hypothetical prediction. Taking the United Kingdom as an example, the case count predicted under no interventions quickly exceeds 10,000 new cases per day, while the observed number ranges below 10,000 per day. Fig. 5 also allows to compare the daily estimated number of new cases from our full model (based on the NPIs that the country actually implemented) with the observed (documented) number of new cases. Here an acceptable degree of agreement is observed for the UK, US, and other countries (Figures for all other countries are listed in Appendix G). Note that these figures solely serve as a visualization of the estimated effect of NPIs on new cases (and credible intervals are thus omitted in these figures).

**Fig. 5.**
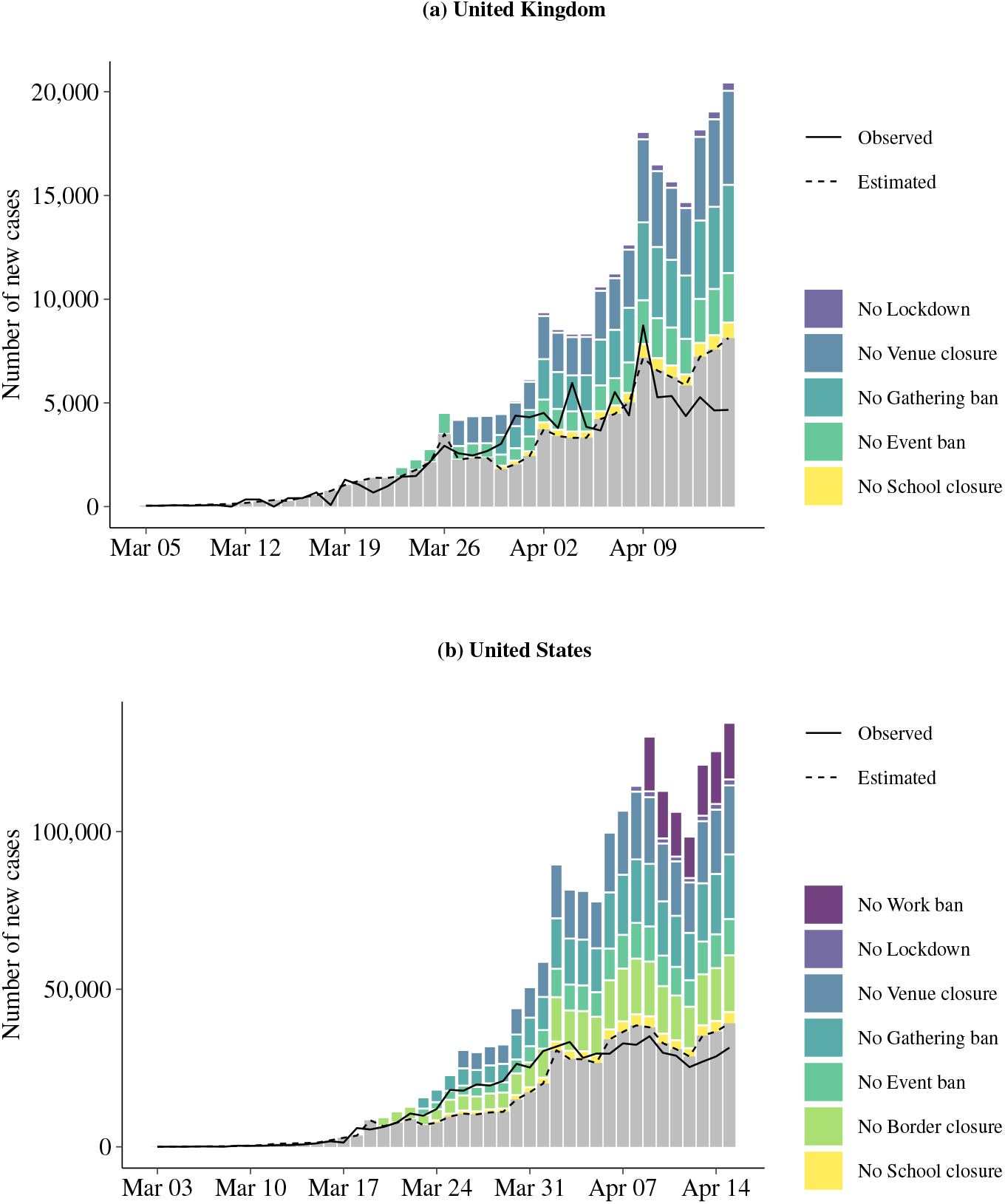
Hypothetical number of daily new cases if non-pharmaceutical interventions (NPIs) were not implemented. The stacked bars illustrate how the mean estimated number of new cases would have changed if NPIs were *not* enforced. The solid line refers to the observed (documented) and the dashed line to the estimated number of new cases. The NPIs are shown for the time period in which they were implemented while considering that their effect is subject to a time delay of 7 days.

## 4. Discussion

A cross-country analysis based on *n* = 20 countries was performed in order to assess the relative effectiveness of NPIs. Four of our seven NPIs were associated with relative reductions in the number of new cases of 30 % or above and a range of credible effects above 15 %, indicating a highly relevant role. Both venue closures and gathering bans were among these four NPIs, and each was implemented by 19 out of *n* = 20 countries in our sample. This is an interesting addition to prior literature, in which these NPIs have received rather little attention and thus deserve further investigation. Work bans also belonged to the group of effective measures, but this NPI was implemented by only six countries, which implied greater uncertainty about the estimate. The NPI border closures was the last member of this group. Simulations confirmed that a rapid dissemination of SARS-CoV-2 was associated with large numbers of undocumented infections [6], especially if not prevented by travel restrictions [8]. However, this measure should be interpreted with caution as it primarily targets international transmissions, which might be more pronounced in early stages of an outbreak as studied in this analysis. In later stages, the effect might be substantially smaller, with this measure primarily steering international travelers or cross-border commuters towards increased social distancing.

School closures restrict access to education with unwanted implications for members of society who are already underprivileged. In our analysis, this NPI belonged to those with only a rather moderate effect estimate, amounting to a 10 % reduction. This finding is in line with prior literature in which the transmissibility of SARS-CoV-2 among children is regarded as comparatively small [11, 23]. The preceding argument also provides a possible explanation for why our model attributed only a small reduction of cases to this NPI. Lockdowns also had only a very moderate and possibly negligible effect. As our effect estimates describe the additional impact of an NPI on top of the other NPIs, the moderate effect of lockdown may be explainable by event bans, venue closures and gathering bans catching already a substantial part of the impact of a lockdown. Summing up the effects of all four NPIs, we obtain in any case a substantial effect. Event bans were located between the groups of NPIs with large and small relative reductions, with an estimated relative reduction of about 25 %, but a wide range of credible effects.

Due to the concurring introduction of NPIs in many countries, it was difficult to distinguish between the effects of single NPIs. This is reflected by large credible intervals and a negative association between effects (Appendix D.2), suggesting that the effect of one NPI may be attributed partially to another. However, with respect to the ranking of NPIs, we were able to demonstrate that we can be fairly confident that venue closures, gathering bans, border closures, and work bans are among the four most effective NPIs and school closures and lockdowns among the two least effective when applied as a combination of NPIs.

Our analysis is subject to limitations. First, our analysis is limited by the type of data utilized and by the need to make modeling assumptions. Using the number of documented cases implies that documentation practices may have an influence on the results. In particular, definitions and documentation practices differed between countries and over time. However, since the rate of new cases was investigated, many deviations from an optimal documentation practice will affect both the number of new cases and the number of existing cases in a similar manner and may thus cancel out. Short-term fluctuations were considered by including day-of-the-week effects and allowing for overdispersion. Moreover, it was taken into account that countries had to develop their documentation practices by starting with at least 100 cases for each country. However, there may still remain an undue influence of country-specific changes in documentation practices over time.

Second, a fundamental issue with our modeling approach is the implicit assumption that any deviation from a constant rate of new cases is explained by the NPIs. It is possible that additional measures or an increasing general awareness encouraged social distancing and hence lead to less infections. If this is the case, such effects will erroneously be assigned to the NPIs and possibly overstate their overall impact.

Third, we could not follow the more correct approach to relate the number of new cases to the number of non-recovered cases, as reliable information on the number of recovered cases was not available. However, within our study period, the number of recovered patients was comparatively low relative to the overall population and, hence, it was justified from our perspective to neglect this issue when analyzing the early stages of the outbreak. With respect to the true infection rate, theoretical simulations [22] suggested a stationary rate in the absence of NPIs for this early phase and, hence, independence from natural attrition phenomena.

Fourth, our modeling assumptions do not account for any interaction between countries and the effect of NPIs. The simplifying assumption of a common effect reflects our aim to estimate an “average” effect and the current lack of power to investigate cross-country variation.

In conclusion, NPIs lead to a strong overall reduction in the number of new documented cases of COVID-19. Venue closures, gathering bans, border closures, and work bans had a high impact, event bans a slightly less pronounced impact, and school closures and lockdowns had a comparably small impact. Our effect estimates provide first and timely estimates to inform the current debate. They should be updated and refined in the future. Additional information may arise from the federal structure of some countries (e.g., the US and Germany), from more detailed case data (e.g., recovery status or age), or from rescinding NPIs.

## Acknowledgements

We acknowledge feedback from the ETH COVID-19 Task Force.

## Data availability

We collected data from publicly available data sources (Johns Hopkins Coronavirus Resource Center for epidemiological data; news reports and government resources for policy measures). All the public health information that we used is documented in the main text, the extended data and supplementary tables. A preprocessed data file is available with the codes.

## Code availability

With publication, codes that support the findings of this study are available from https://github.com/mis-research/covid19_npi_effectiveness.

## Contributions

NB contributed to conceptualization, data collection, data analysis, results interpretation and manuscript writing. EvW contributed to data collection, data analysis and manuscript writing. AS contributed to data analysis and manuscript writing. BK, AC, PB, and JPS contributed to data collection. DT contributed to results interpretation. WV contributed to conceptualization, data analysis, results interpretation and manuscript writing. SF contributed to conceptualization, results interpretation and manuscript writing.

